# Digitalization impacts the COVID-19 pandemic and the stringency of government measures

**DOI:** 10.1101/2022.06.27.22276377

**Authors:** Helen Heinrichs, Florian Mueller, Lucia Rohfleisch, Volkmar Schulz, Steven R. Talbot, Fabian Kiessling

## Abstract

COVID-19 poses a significant burden to populations worldwide. Although the pandemic has accelerated digital transformation, little is known about the influence of digitalization on pandemic developments. Therefore, this country-level study aims to explore the impact of digital adoption on COVID-19 outcomes and government measures. Using the Digital Adoption Index (DAI), we examined the association between countries’ digital preparedness levels and COVID-19 cases, deaths, and stringency indices (SI) of government measures during the early phase of the pandemic. Gradient Tree Boosting pinpointed essential features related to COVID-19 trends, such as digital adoption, populations’ smoker fraction, age, and poverty. Subsequently, regression analyses indicated that higher DAI was associated with significant declines in new cases (β=-362.25/pm; *p*<0.001) and attributed deaths (β=-5.53/pm; *p*<0.001) months after the peak. When plotting DAI against the SI normalized for the starting day, countries with higher DAI adopted slightly more stringent government measures (β=4.86; *p*<0.01). Finally, a scoping review identified 70 publications providing valuable arguments for our findings. Digital adoption shows a positive trend in handling the current pandemic and facilitates the implementation of more decisive governmental measures. Improving the distribution of digital adoption may have the potential to attenuate the impact of COVID-19 cases and deaths.

## Introduction

The global coronavirus disease 2019 (COVID-19) outbreak has kept the world on hold for some time. Countries have suffered large numbers of infections and deaths ^1,2^, which tremendously affected economies and societies^3,4^. As a response, governments have implemented coordinated containment and mitigation strategies ^5^. Although authorities worldwide have employed similar policies, such as work and school closures, border restrictions, quarantine, or lockdown, their impact on the pandemic and the stringency of implementation have varied across countries ^6-8^. Recent research suggests that the effectiveness of policies depends on many factors, including socioeconomic and demographic conditions as well as cultural and political structures ^5,9^. In addition, digital information technology may be a distinguishing factor in determining the course of the pandemic ^10^.

The pandemic has accelerated digital transformation in many areas of life. The health care sector is rapidly adopting telemedicine ^10^, schools are working with collaborative online platforms ^11^, and business models are becoming more hybrid ^12^. However, little is known about how digitalization has shaped the crisis and how it has been managed. Asian countries have introduced many innovative technologies to attenuate the spread of COVID-19. South Korea and Singapore strengthened their digital contact tracing tools by promptly implementing surveillance technologies ^13,14^. Hong Kong and Taiwan used government-issued mobile phones and wristbands as quarantine compliance measures ^10,15^. Such measures are potentially relevant factors in controlling the pandemic, leading to less stringent governmental measures ^8,13^. The effectiveness of digital contact tracing in controlling outbreaks has been discussed in much earlier work by Fraser et al. ^16^ and has recently been adapted to the case of COVID-19 ^17^. However, it is unclear how digital tracing and surveillance are implementable in regions with diverse digital infrastructure. Evidence in existing research focuses on the promises of digitalization. Nevertheless, as described in previous papers, contact tracing requires extensive digital infrastructure ^13-15^. The pandemic has exposed fundamental gaps in the universality of digital access, resulting in challenges in the implementation and use of digital technologies ^18^.

This study aimed to explore the association between digitalization and COVID-19 developments at the country level. Specifically, we examined whether a country’s level of digital adoption prior to the pandemic may have influenced the number of COVID-19 cases, deaths, and the stringency index (SI) of government actions. Our approach allows for evaluating the impact of digital adoption on pandemic propagation and government activity using an international database. The findings highlight the need for efficient and technologically advanced infrastructures to combat the current and future potential health crises.

## Results

We retrieved data from the open-access COVID-19 dataset, available via GitHub and maintained by *Our World in Data* ^19^. These data are updated daily and facilitate an easy-to-use and comparative framework for this country-level study. It also entails the SI from the Oxford COVID-19 Government Response Tracker, which we included as a standardized method for comparing the stringency of government measures across countries ^20^. We used the reported data until March 10, 2021, to focus on the early phase of COVID-19, when countries took initial actions and new technological tools were not yet available or being developed to assess countries’ digital preparedness facing COVID-19. The World Bank’s DAI expresses countries’ levels of digital adoption across three dimensions of the economy: government, business, and people. ^21,22^. Each of the three sub-indices comprises various equally weighted and normalized indicators on a 0 to 1 scale, while the DAI represents the simple average. The “government” pillar combines online public services, digital identification, and core administrative systems. The “business” pillar includes the number of secure servers (in millions), internet speed (kbps), percentage of businesses with websites, and 3G coverage. Finally, the “people” pillar indicates mobile and internet access at home. The most recent year for which DAI country data was available was 2016.

### The more advanced countries’ digital adoption, the lower the number of cases and the faster new cases decline

Gradient Tree Boosting (GTB) was first applied to compare DAI and other demographic and health-related country-specific variables with total COVID-19 cases in 2020. At the peak of COVID-19 cases, GTB showed a strong dependence on digitalization across countries (0.23, SD±0.12) next to the proportion of female smokers (0.32, SD±0.22) and older people (0.22, SD±0.14). Male smokers (0.14, SD±0.04) and per capita gross domestic (0.09, SD±0.05) were also essential characteristics (Fig. 2a). Values in parentheses indicate the relative feature importance and standard deviation.

**Figure 1.**
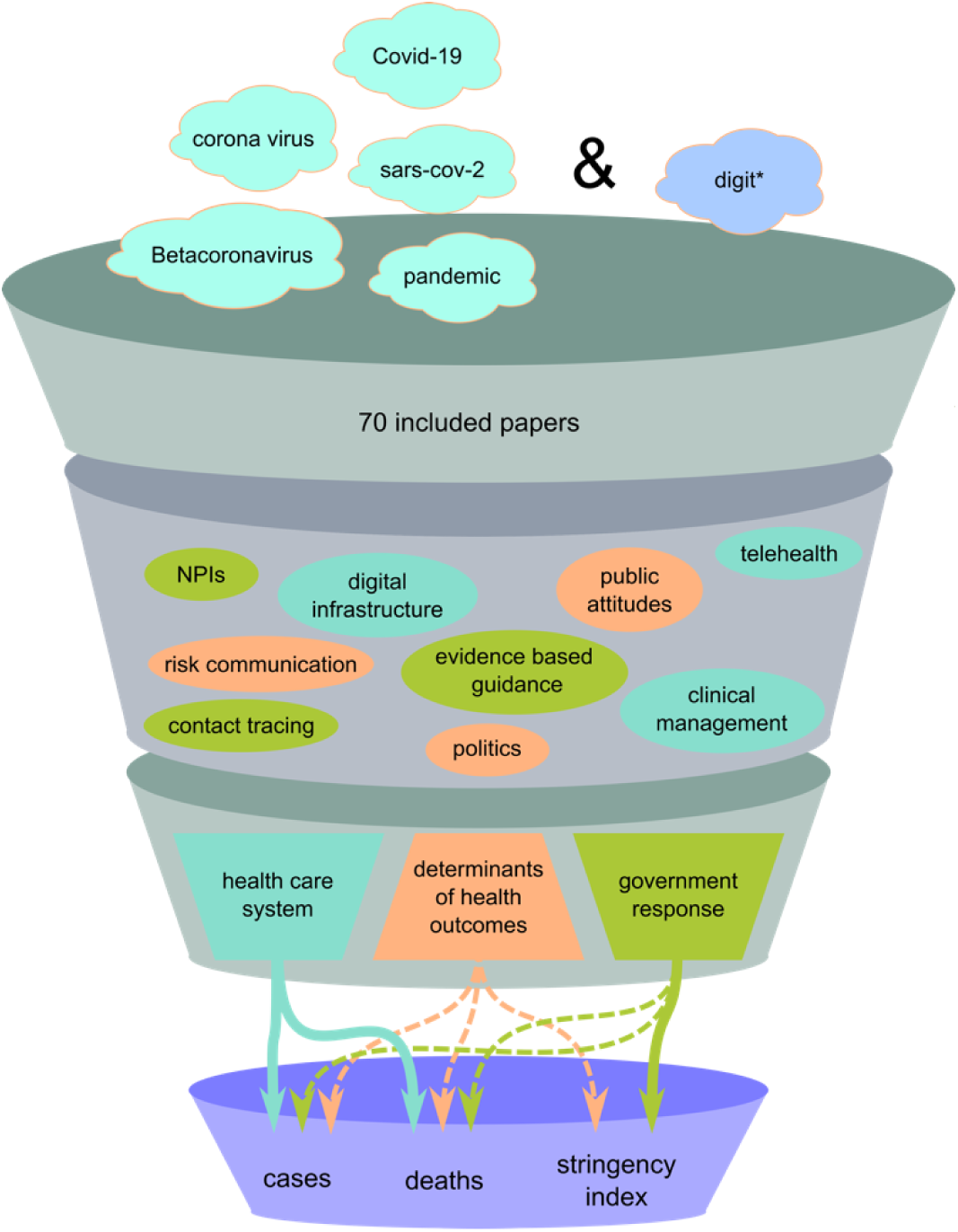
Funnel model of included themes and keywords in the scoping review. The funnel model illustrates the scoping review process. We began our literature review with a broad search strategy that included COVID-19-related terms (e.g., “Covid-19” OR “sars-cov-2” OR “pandemic”) and digit*, which represents the digital domain with different word endings (as indicated by an asterisk (*)). After an extensive selection process, 70 papers were included in our review. From these papers, we identified keywords reflecting recurring aspects of the literature. These were grouped into the following themes: i) health care system, ii) government responses, and iii) determinants facilitating or hindering public health outcomes. Eventually, the scoping review offered essential arguments for our statistical findings. (Note: NPIs = non-pharmaceutical interventions; NPIs are public health measures to prevent and/or mitigate transmission of SARS-CoV-2 that do not rely on medications or vaccination. These include, but are not limited to, surveillance measures, travel restrictions, and recommended use of face masks ^23^).

**Figure 2.**
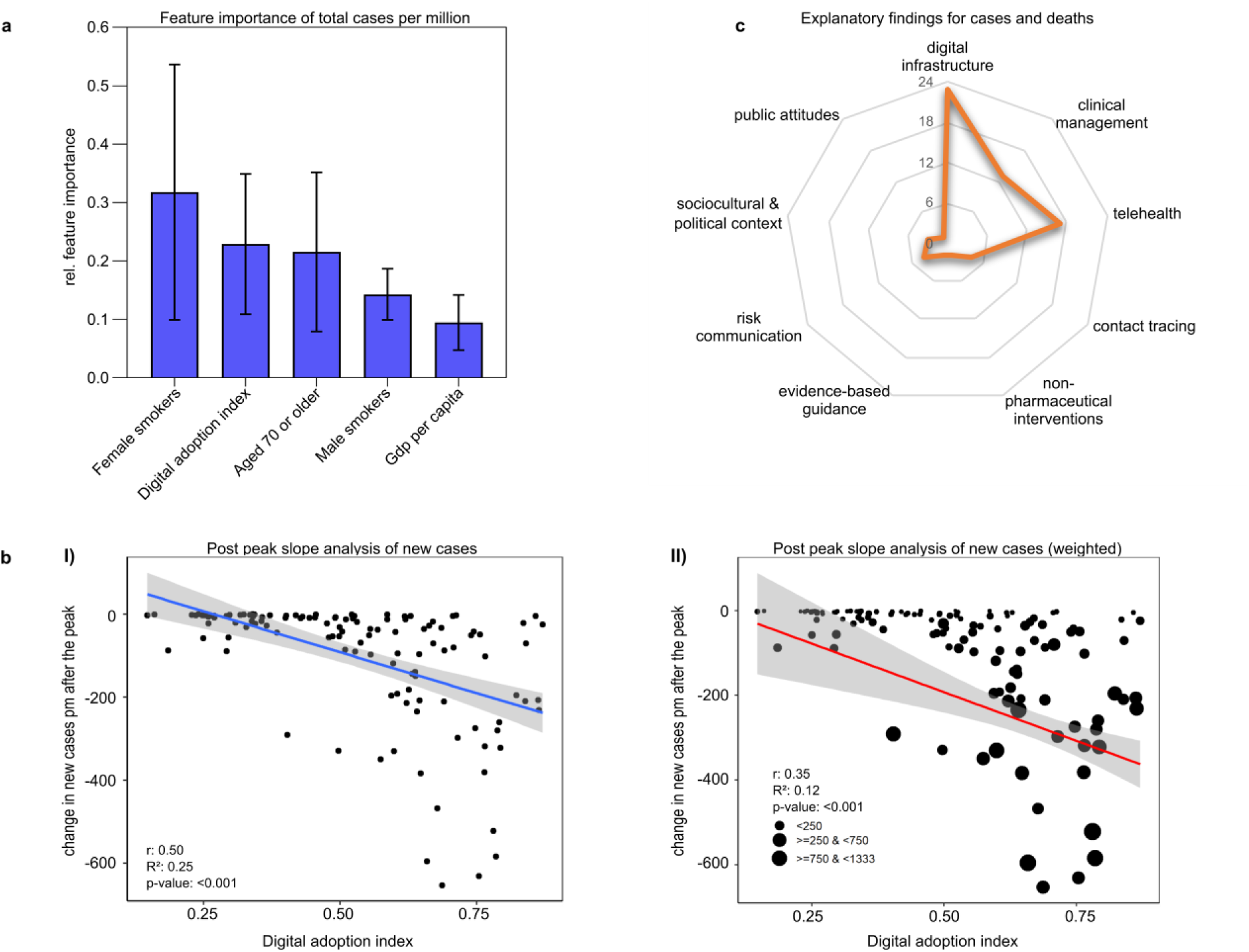
The impact of digital adoption on the development of COVID-19 cases. **a** The gradient tree boosting results show the five most important characteristics for peak total COVID-19 cases in 2020 out of 15 features. Relative feature importance was normalized, and the sum was set to 1.0. Data are expressed as mean ± standard deviation. **b** Change in new cases pm after the peak in 2020-2021. I) The result of the simple linear regression illustrates the effect of the DAI on the reduction in new COVID-19 cases (7-day-smoothed) pm in the post-peak period. Countries with higher DAI were more efficient in significantly reducing the number of new cases. II) The second regression is weighted by the delta of minimum and maximum new cases to show the impact of more successful countries in lowering the new case numbers. The blue (I) and red (II) lines indicate the linear fits of regression analyses. The areas of the 95% confidence intervals are shaded gray. DAI is expressed as an arbitrary unit. **c** The radar chart displays the keywords addressed in the “health care system” theme of the scoping review. The scale for the radial axis diagrams shows how frequently the keywords occurred in the included studies. Papers may contain different numbers of keywords.

Using linear regression, we plotted the change in new cases per million people (pm) by countries’ peaks against DAI. We scrutinized the slope to determine how DAI potentially counteracted pandemic trends within months after reaching maximum new cases. Figure 2b shows a significant decline in new COVID-19 cases with increasing DAI in most countries, trending toward 362 fewer new cases per unit DAI increase (β=*-*362.25/pm, *p*<0.001). This trend analysis was repeated with a regression weighted by the difference in caseload, also showing a greater ability to reduce new cases pm with increasing DAI (β=*-*446.54/pm, *p*<0.001) (Table 1).

**Table 1.** Linear regression analyses of the association between DAI and changes in COVID-19 new cases and deaths months after their peaks.

| | $\beta$ | Std. Error of $\beta$ | t | p-value |
| --- | --- | --- | --- | --- |
| **Effect of DAI on changes in new cases (smoothed) pm** |  |  |  |  |
| Constant | 92.9 | 30.46 | 3.049 | 0.003* |
| DAI | -362.25 | 53.79 | -6.734 | 0.001** |
| **Effect of DAI on changes in new cases (smoothed) pm (weighted)** |  |  |  |  |
| Constant | 35.43 | 68.46 | 0.518 | 0.606 |
| DAI | -446.54 | 101.06 | -4.419 | 0.001** |
| **Effect of DAI on changes in new deaths (smoothed) pm** |  |  |  |  |
| Constant | 1.02 | 0.73 | 1.398 | 0.164 |
| DAI | -5.53 | 1.29 | -4.293 | 0.001** |
| **Effect of DAI on changes in new deaths (smoothed) pm (weighted)** |  |  |  |  |
| Constant | -3.42 | 1.97 | -1.738 | 0.084 |
| DAI | -3.79 | 2.99 | -1.269 | 0.207 |

\* $p \leq 0.05$ ; \*\* $p \leq 0.001$
Notes: The regression coefficient ( $\beta$ ) is the degree of change in the outcome variable for every one-unit change in the predictor variable. The t-statistic is the regression coefficient divided by its standard error. Digital adoption is expressed in arbitrary units of DAI.

Of the 5012 articles retrieved by our search, we removed 1454 duplicates and another 3450 records during the title and abstract screening. Finally, of the 108 records eligible for the full-text screening, 70 met the inclusion criteria and were included in the scoping review (Supplementary Fig. 3). We grouped keywords into the following themes: i) health care system, ii) government responses, and iii) determinants facilitating or hindering public health outcomes (Fig. 1). In this results section, we discuss the most prominent findings related to the first two themes, and in the Supplementary Results section, we discuss the determinants of public health outcomes (Supplementary Fig. 6). Many articles on digital technologies covered cases and deaths, summarized under “health care system”. Supplementary Fig. 4 and Supplementary Fig. 5 provide some descriptive findings on the geographical mapping of included records.

### Digital technologies within the health care system

The scoping review’s first theme (n=26) addressed health care technological features that may affect care-delivery processes during outbreaks or that were present before the pandemic (Fig. 2c). Telehealth involves various technical methods (e.g., virtual consultations or remote patient monitoring) rapidly adopted in the US, UK, Australia, and elsewhere ^24-26^. Such electronic communication tools improve access to care by eliminating the additional risk of exposure ^24^. With resource shortages looming, Scott et al. introduced the National Emergency Tele-Critical Care Network to deliver care and expertise via mobile devices across the US ^27^. Chatterjee et al. provided an overview of integrated communication systems in South America ^25^. Ecuador, Bolivia, and Peru, which score medium in DAI, introduced telemedicine platforms providing access to specialists virtually. Chile, achieving a higher DAI, enforced a new model of digital hospitals with upcoming features like electronic health records (EHRs). Additionally, they propose using IoT or AI-based devices to assess health parameters more accurately ^25^. Wood et al. investigated the state EHRs in the UK ^28^. While researchers initially lacked access to critical population health data, the British Heart Foundation Data Science Centre developed a digital environment with secure access to crucial COVID-19 data for researchers. During the COVID-19 outbreak, China rapidly expanded the establishment of internet hospitals. These combine various web-based services for the public, such as telemedicine, drug delivery, and psychological counseling ^29,30^. An early adopter of robots, Guangdong Provincial People’s Hospital assisted the medical staff by disseminating health information or delivering medicines to isolation wards ^31^. While telecommunication facilitates medical treatment in First Nations or sparsely populated islands ^26^, illiterate or low-income populations are severely affected by the digital divide and face unequal access to telehealth ^25^. Despite a high mobile penetration in India, the introduction of eHealth services proved difficult for similar reasons ^26^.

### The more advanced countries’ digital adoption, the lower the number of deaths and the faster new deaths decline

We employed GTB to identify essential features of the peak numbers of deaths pm (Fig. 3a). We found that the proportion of female smokers (0.49, SD±0.12) held the highest feature importance, followed by extreme poverty (0.17, SD±0.02) and the population aged 70 or older (0.11, SD±0.07). Additionally, GTB indicated feature importance for DAI (0.13, SD±0.05) and the proportion of male smokers (0.1, SD±0.03).

**Figure 3.**
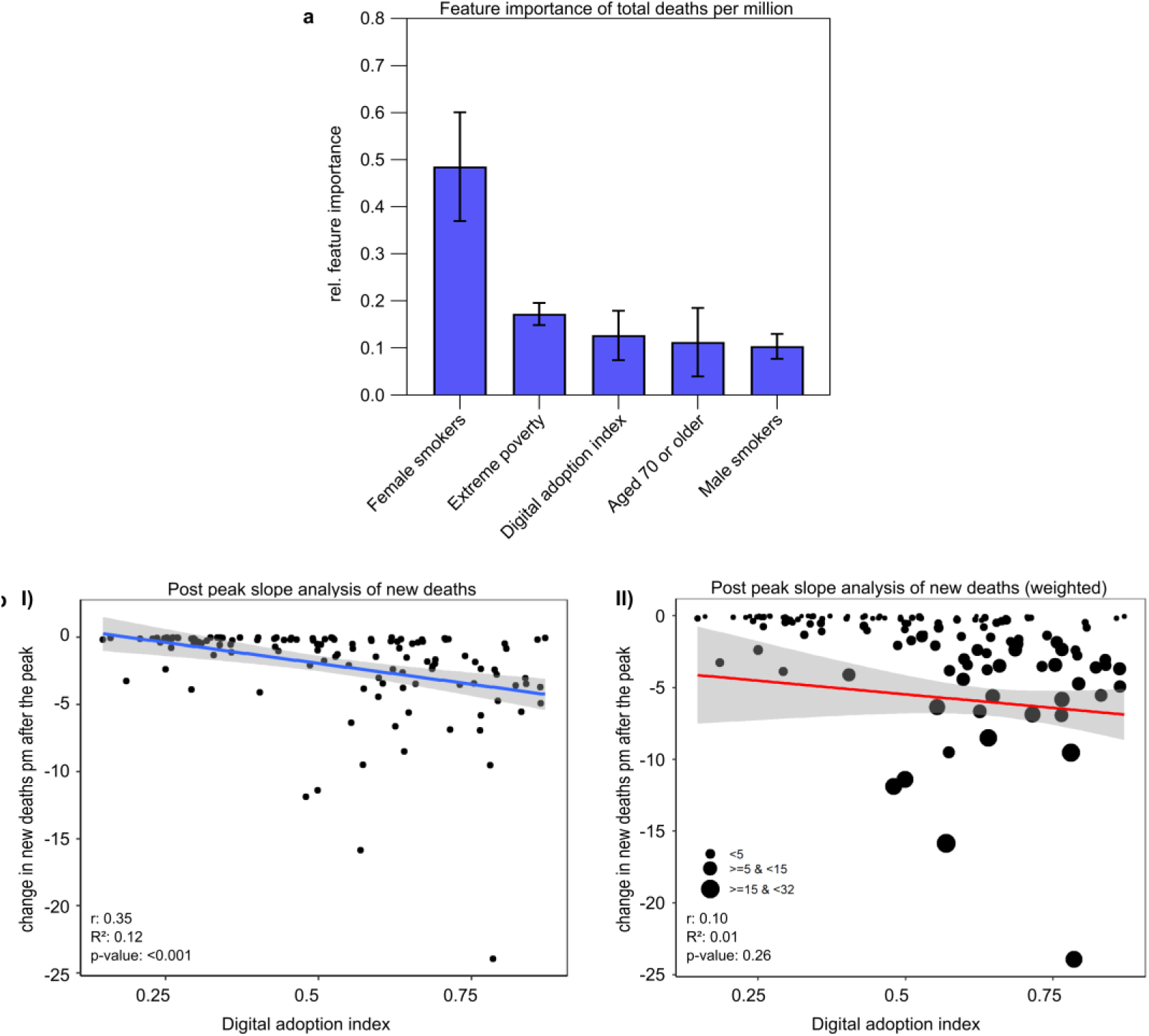
The impact of digital adoption on the development of COVID-19 deaths. **a** Gradient tree boosting results show the top five important features for peak COVID-19 deaths in 2020. Relative feature importance was normalized, and the sum was set to 1.0. Data are expressed as mean ± standard deviation. **b** Regression results show the effect of the DAI on reducing new deaths pm during the post-peak period in 2020-2021. I) Countries with higher DAI could significantly reduce their number of new COVID-19 deaths (7-day-smoothed) months after the peak. II) In the weighted regression analysis, the significant effect of the DAI on reducing deaths is no longer visible. The blue (I) and red (II) lines indicate the linear fits of regression analyses. The area of the 95% confidence interval is shaded gray. DAI is expressed as an arbitrary unit.

Regression analysis shows the impact of digitalization on post-peak COVID-19 deaths (Table 1; Fig. 3b). Higher DAI was associated with a significant decrease in new deaths pm, resulting in approximately 5.53 (*p*<0.001) fewer deaths pm after the peak in 2020/2021. The slope expresses the decline in cases after reaching the peak for each country. We performed another regression analysis weighted by the decrease in the number of deaths, showing the same trend (β=-3.79/pm; p=0.207).

### The influence of digitalization on the stringency of government measures

The most important features identified with GTB and associated with SI were hospital beds per thousand people (0.24, SD±0.03), followed by DAI (0.22, SD±0.03). Other informative features included extreme poverty (0.19, SD±0.04), male smokers (0.17, SD±0.02), and diabetes prevalence (0.17, SD±0.02) (Fig. 4a).

**Figure 4.**
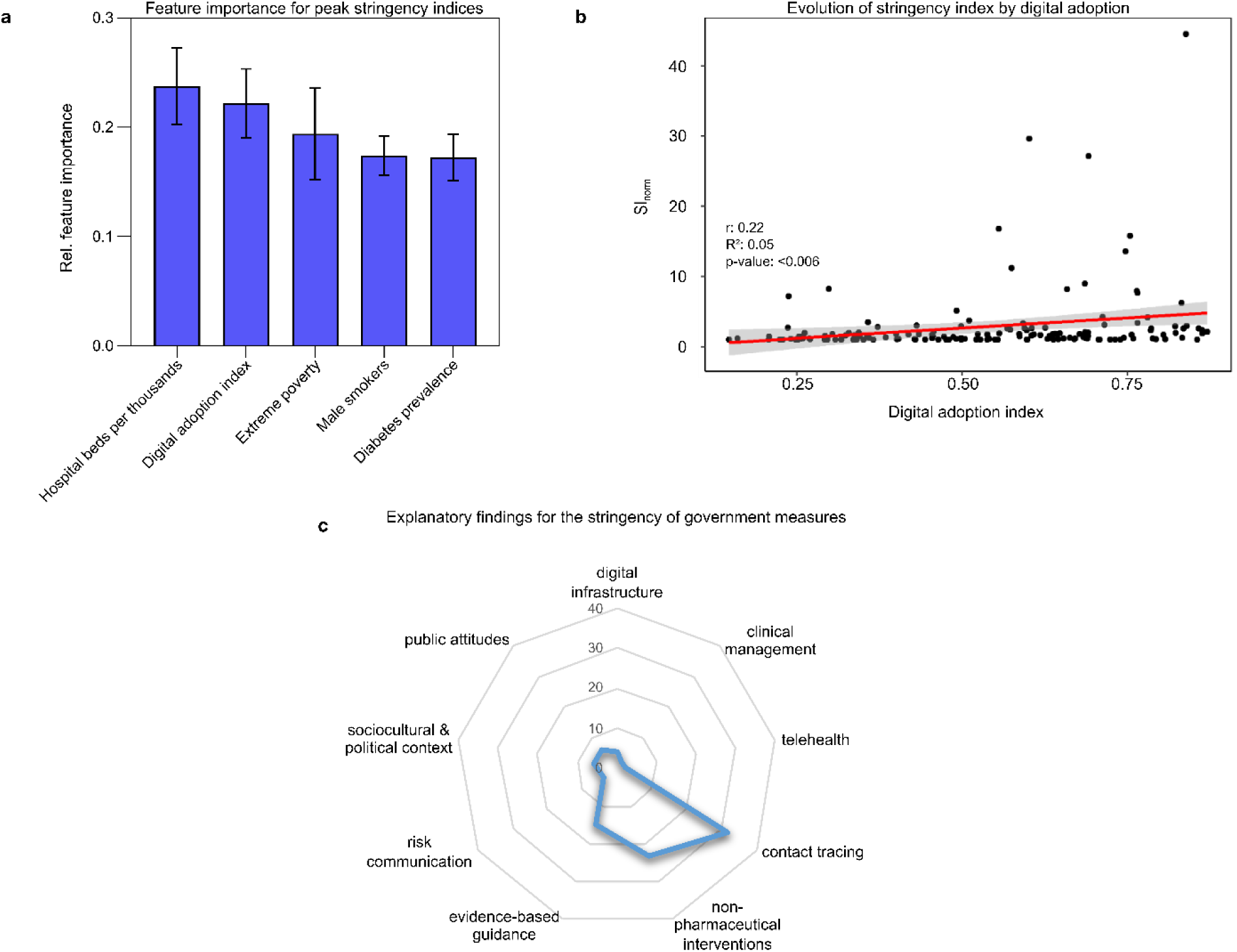
The impact of digital adoption on the stringency of government measures. **a** GTB shows the relative importance of characteristics on government stringency indices. Relative feature importance was normalized, and the sum was set to 1.0. Data are expressed as mean ± standard deviation. **b** The regression model shows that, on average, countries with a higher DAI adopted slightly more stringent policy measures during the first year of the pandemic. To allow for a suitable comparison of the stringency index across countries, we adjusted for the different onsets of the SI (SInorm= SImean/SIinitial). The area of the 95% confidence interval is shaded gray. **c** Findings related to government responses within the scoping review: contact tracing, non-pharmaceutical interventions, and evidence-based guidance. The scale for the radial axis diagrams shows how frequently the keywords occurred in the included studies.

The final regression model shows that, on average, countries with a higher DAI adopted slightly stricter policy measures during the first year of the pandemic (β=4.86/pm, *p*<0.01) (Fig. 4b). After plotting DAI against mean SI, we identified three trends in the data and grouped the DAI accordingly. Then, the estimated effect of each group (low, medium, and high DAI) was analyzed by including the DAI:class interaction term in the model. Subsequent regression analyses of all three classes showed that the interaction of DAI and its high class (>0.68) could significantly reduce the mean SI compared to the interaction of DAI and low class (β=-55.26, *p*<0.05) (Table 2). See Supplementary Table 1 and Supplementary Fig. 2 for more information on the classification of DAI classes.

**Table 2.** Linear regression models of the overall effect of DAI on SI and its regression per class (low, medium, high).

| | $\beta$ | Std. Error of $\beta$ | t | p-value |
| --- | --- | --- | --- | --- |
| **Overall effect of DAI on SI<sub>norm</sub>** |  |  |  |  |
| Constant | -0.40 | 1.00 | -0.398 | 0.691 |
| DAI | 4.86 | 1.75 | 2.780 | 0.006** |
| **Effect of low, medium, and high DAI on the trend in SI** |  |  |  |  |
| Constant | 38.61 | 12.10 | 3.191 | 0.003** |
| low [0;0.36] | 54.18 | 42.01 | 1.290 | 0.205 |
| Constant | 71.18 | 9.98 | 7.133 | 0.001** |
| medium [0.37;0.68] | -10.61 | 18.23 | -0.582 | 0.562 |
| Constant | 102.36 | 18.75 | 5.458 | 0.001** |
| high [0.69;1] | -55.26 | 24.32 | -2.272 | 0.028* |

\* $p<0.05$ ; \*\* $p<0.001$
Notes: The regression coefficient ( $\beta$ ) is the degree of change in the outcome variable for every one-unit change in the predictor variable. The t-statistic is the regression coefficient divided by its standard error. Digital adoption is expressed in arbitrary units of DAI. The numbers in squared brackets indicate the intervals of the classes.

### Digital technologies in government responses

46 of 70 review articles offered insights into government responses (Fig. 4c). Throughout the pandemic, contact tracing and surveillance management were utilized to support government decision-making ^32,33^. Southeast Asia led the way in leveraging digital technologies to track transmission routes early in the pandemic. Digital contact tracing in South Korea, Singapore, and Hong Kong ^34,35^ is attributed to the successful containment of COVID-19 in its early stages ^15^. The countries’ populations are particularly obedient to following government guidelines and using their cell phones’ GPS. Israel also collected cell phone location data to aid contact tracing ^36^, and Taiwan ordered staying-at-home measures immediately after detecting a case ^34^. Some Asian countries have been exceptionally proactive and diligent in integrating digital governance. A case study in Wuhan ^32^ exhibited community-based digital contact tracing, such as the Health QR Code for smartphones, a few weeks into the pandemic. This QR code held personal and health information and was regularly checked by contact tracers to monitor people’s mobility. The government used this to divide Wuhan’s inhabitants into community grids according to their risk status. Real-time data enabled governments to identify clusters of viral spreading, eventually reducing total infections and hospitalizations shortly after implementing the app. In Bangladesh, the government initiated mobile awareness initiatives where people received preventive information on their cell phones if they were subscribed to a phone operator ^37^. While some digitalized countries have been more proactive, Germany and Austria were relatively slow to respond to the pandemic and eventually introduced stringent measures ^35^. The initiation of lockdowns or travel restrictions in the US appeared less organized and varied between states ^15^. Other studies show great potential for implementing transmission models and providing evidence-based guidance to governments ^38^. For example, Leung et al. established a framework to track Hong Kong’s replication number of COVID-19 in near real-time, without multi-day delays, using age-specific digital mobility data ^39^. Cencetti et al. introduced a model for contact tracing combined with direct quarantine orders for infected individuals ^40^.

To identify potential confounding characteristics for DAI, we performed an ANOVA with countries’ income levels. ANOVA followed by a Games-Howell post-hoc test revealed significant differences among all four income levels, *F*(3,160)=154.87, *p*<0.001, and showed that DAI was proportionally higher in high-income countries (*p*<0.001) (Fig. 5).

**Figure 5.**
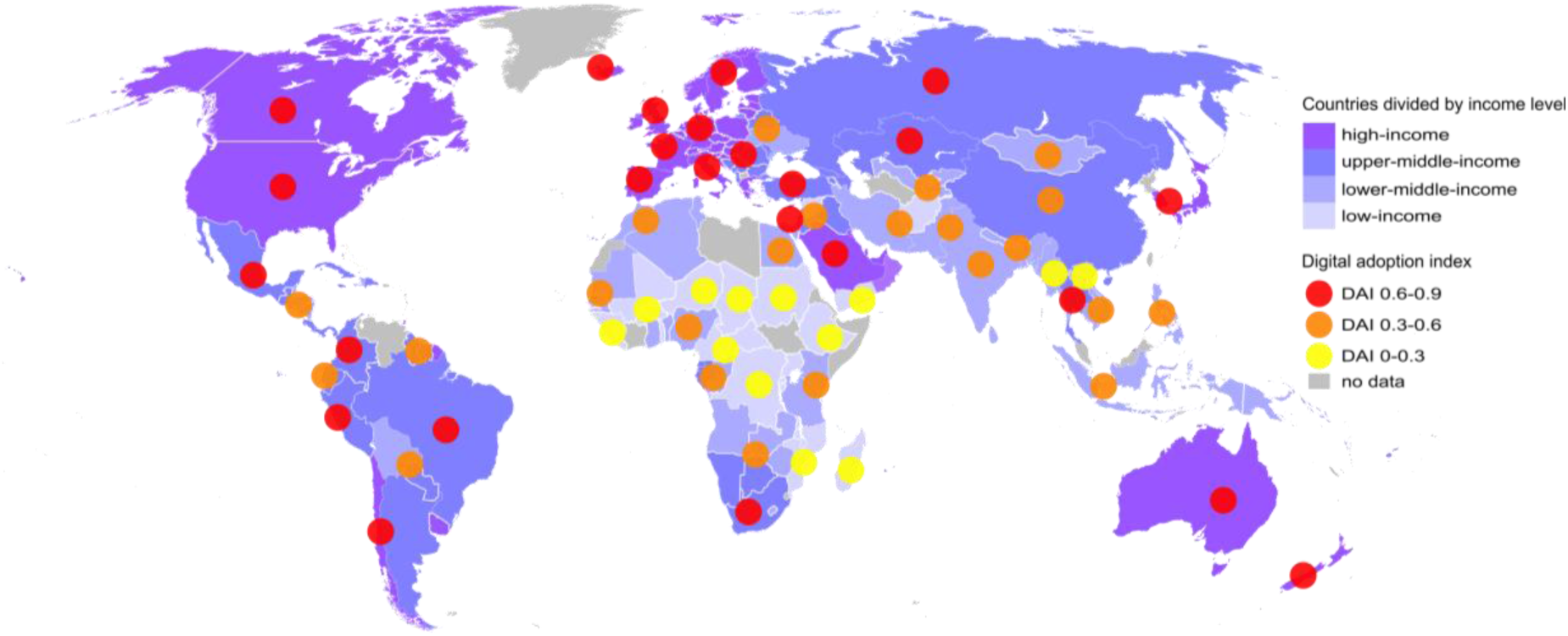
Global level of digital adoption by income. Digital adoption by country is represented by the digital adoption index (DAI), chosen as the one representative score covering numerous countries. The DAI is based on three sub-indices implying governments, businesses, and people. Regions include economies at all income levels ^41^. Adoption of digital technologies varies across countries, with higher- and upper-middle-income countries scoring a higher DAI. As Singapore reached the highest DAI index of 0.87, we divided the DAI into three increments up to 0.9.

## Discussion

COVID-19 prompted the rapid and enthusiastic implementation of numerous digital technologies and processes. Given the ongoing devastating situation in many countries and the risk of further SARS-CoV-2 transmission, optimizing digital transformation is necessary but requires fundamental digital adoption. Our findings show that digitalization will not act as a single silver bullet to prevent outbreaks but that digital adoption significantly reduces cases and deaths and influences governments’ SI. Therefore, a country’s digital adoption level may be a determinant in mitigating COVID-19 and future pandemics.

GTB results indicate associations between several variables, including DAI and pandemic trends. DAI was found in this study to be associated with the three outcome variables: cases, deaths, and countries’ stringency indices. Furthermore, COVID-19 cases and deaths are linked to populations’ proportion of smokers and elderly. Some studies attribute smoking to a worse outcome after contracting the virus ^42-44^, similar to how it was a risk factor for mortality in MERS-CoV cases in South Korea in 2015 ^45^. The risk of death rises with age ^43,46^ and is highest in countries with elderly populations ^47^. Also, the risk of comorbidities, e.g., diabetes, increases with age, which increases the likelihood of a severe infection course ^48^. COVID-19 cases were associated with lower per capita gross domestic ^49^ and deaths with poverty ^50^. A possible explanation for the association between hospital bed capacity and SI is the increased pressure on hospital beds, which stimulated governments to introduce new policies ^51^.

Linear regression analyses show the relationship between DAI and the evolution of the pandemic. Countries with higher DAI proved more resilient in managing cases and deaths than countries with lower DAI. Jurzik et al. note that higher levels of digital adoption likely contributed to the ability of some Asian governments to respond effectively, highlighting the digital divide ^52^. The COVID-19 pandemic has exacerbated the digital divide, disproportionally affecting minorities ^53^ and people living in relatively poorer regions ^54^. We found similar results. In particular, we found a positive association between DAI and country income levels, with countries with higher digital adoption also being among the higher-income groups. WHO’s “Global Strategy for Digital Health 2020-2025” advocates for accelerated implementation and universal and equitable access to digital health and health policies ^55^. Digital health, including competencies in communication and information technologies, internet, and broadband access, is classified as a determinant of health ^56,57^. A lack of digital literacy and skills can put people at risk for adverse health outcomes ^58^.

Review papers explored digital technologies and tools used during the pandemic. Additionally, some studies underline that sociocultural and political contexts, and public attitudes can strongly influence the effectiveness of government measures. Although countries with higher DAI have better prerequisites to cope with pandemics, countries like the US ^36^ have failed to promptly adopt contact tracing mainly because of privacy concerns ^40^. Knowledge of previous respiratory syndrome outbreaks was invaluable for effective early response in Singapore, Taiwan, and Hong Kong ^59^. After the MERS-CoV outbreak, South Korea implemented several tracing methods allowing authorities to collect data on peoples’ mobility via credit cards and cell phones ^60^. These approaches require numerous contact tracers, but affluent Western countries lacked such positions when COVID-19 struck^61^. However, the success of surveillance management in Asian countries comes at a price: a lack of privacy and freedom ^62^. Some of the countries scoring high in DAI, like England, France, Germany, and the US, were more reluctant to use digital measures and introduced restrictive non-pharmaceutical interventions (NPIs), e.g., lockdowns. These had farer-reaching negative personal, social, and economic impacts ^62^. For NPIs to contribute meaningfully to the containment of COVID-19, they should be used effectively and targeted ^23^. Countries that deployed digital surveillance and contact tracing remained pioneers in managing disease burden ^10^, ultimately strengthening people’s autonomy to maintain near-normal lives ^62^. However, not all digital innovations are transferable to countries struggling with massive outbreaks. Internet use has spread globally but is less well utilized in poorer countries, posing a barrier to basic requirements for using digital technologies in those countries ^22^.

This study has some limitations due to its country-level ecological study design. The lower fits of regression models suggest inherent variation in the data likely caused by measurable and non-measurable confounding factors such as inaccurate reporting of COVID-19 data or the heterogeneous nature of the pandemic. Therefore, this study cannot explain a causal relationship, but it does show a clear trend between digital adoption and COVID-19 measures. We intentionally selected the country-level data to provide an initial global trend analysis. Future research could consider expanding the covariates to include those identified during GTB, countries’ social, economic, environmental, and health characteristics, and additional information on testing and vaccination. In addition, disaggregating these data into individual-level data would allow for a more in-depth analysis of the determinants of COVID-19 trends and a more targeted approach to mitigating the pandemic’s effects. Finally, the choice of timing may have influenced statistical analyses. The pandemic was marked by its highly evolving nature in data, and that case and death rates changed substantially. Therefore, we took the data at the time of their maximum growth rate and computed the post-peak period for regression analyses, allowing practical and efficient monitoring of pandemic evolution.

These results suggest that scaling up digital adoption might potentially serve as an effective approach to attenuate COVID-19 cases and death rates. To our knowledge, this is the first population-level study to examine the influence of digital adoption on pandemic trends worldwide. In doing so, we identified digital adoption as a determinant that may have contributed to the severity of COVID-19 outcomes and led to a faster decline in new cases and deaths but also resulted in stricter government measures. Thus, based on this manuscript, policymakers may recognize and appreciate the major impact of digitalization on societies and health systems worldwide. Motivated by this study, we encourage future research to conduct in-depth analysis using within-population-level data when available to adapt for other underlying confounding factors.

## Methods

### Feature identification and ranking

GTB is a supervised machine learning algorithm building classification and regression models for labeled data sets (training data) ^63,64^. GTB models are organized as an ensemble of sequential decision trees, evaluated as simple comparisons with binary outcomes. The tree topology allows following the inference process and identifying the most relevant features based on the frequency and position of the used features in the decision trees, referred to as feature importance ^65^. Thus, GTB offers better data interpretability. A sequential elimination method was applied to identify the most relevant input features. The XGBoost implementation handled missing input features intrinsically ^66^. For the identification of important features, we took countries’ DAIs’ and country-level demographics and health-related parameters adapted from the COVID-19 dataset (population density, median age, aged 65+, aged 70+, gross domestic product per capita, extreme poverty, cardiovascular-related death rates, diabetes prevalence, share of female and male smokers, handwashing facilities, hospital beds, life expectancy at birth, and human development index) ^19^. We identified peaks of total confirmed cases and deaths of COVID-19 pm and SIs in 2020 and performed three analyses separately. We conducted a hyperparameter search to limit the complexity of GTB models ^66^. Therefore, a random split into training and validation data was applied. The found set of hyperparameters was fixed for the following steps. We trained GTB models following a 10-fold cross-validation scheme to probe the data repeatably. For every GTB model of the ten training rounds, feature importance values were extracted for all input features and averaged. We discarded the least important feature and repeated the analysis until only one remained. If multiple features showed equal importance, the discarded one was randomly chosen.

### Linear regression analyses

Simple linear regression showed the relationship between DAI and the change in COVID-19 cases and deaths. To express the change in COVID-19 new cases pm and new deaths pm (7-day-smoothed), we determined countries’ post-peak case and death reductions. Starting at peak values for each country of log-transformed data, we fitted linear regressions to the subsequent minimum of monthly maxima between 2020 and 2021. Then, we extracted slope parameters (change of log¬10 (new cases and deaths 7-day smoothed pm) over time) for each country. Results were visualized by plotting DAI against slopes using ggplot2 ^67^. Supplementary Fig. 1 shows how we estimated the decline in new deaths pm in the post-peak period, using a few country examples. The same method was applied to determine the post-peak period in new cases pm. We excluded countries that a) were unable to reduce their death rates, b) experienced an increase in death rates that exceeded half of the peak, or c) reached the peak at the end of our dataset. Based on the defined criteria, the following countries were excluded from assessing the post-peak change in new cases (n=121) and deaths (n=123): Cambodia, Dominica, Laos, Marshall Islands, Saint Kitts and Nevis, Samoa, Solomon Islands, Timor, Vanuatu (n=9). In addition, Hong Kong and Macao were excluded from regression analyses due to missing DAI data. In another step, we added a weight to the post-peak regression analysis by measuring the difference between the previously determined monthly minimum and the maximum of new cases, respectively, deaths (Δ=max-min).

Following this, we analyzed the association between DAI and SI. To allow appropriate comparisons of SIs between countries, we normalized the actual SI by the initial SI (SI_initial_) per country in the dataset (SI_norm_= SI_mean_/SI_initial_). We then performed linear regression between DAI and SI_norm_. Further, when plotting the SI_mean_ against DAI, we noticed three micro-trends in data. To characterize these trends with linear models, DAI was thresholded in the range [0·2: 0·8] in 0·01 steps. The range was chosen so that sufficient data remained to the left of the lower and right of the upper threshold to perform regression. The value of each DAI step was compared to the actual DAI value in each country and classified with a binary function as either “smaller” or “larger” than the current DAI-threshold. This resulted in n=61 labeled data sets with SI_mean_, DAI, and binary class information for each of the 155 countries. We hypothesized that a local change in SI trend would show as a significant change in the statistical interaction of DAI with the binary class. Therefore, each data set was analyzed with a linear regression (SI_mean_ ∼ DAI:class), and the p-value of the interaction was monitored. We observed two breaks in the inferential interaction analysis, where the p-value changed from *p*≤0.05 to *p*≥0.05 (Supplementary Table 1, Supplementary Fig. 2). These thresholds were used to classify the DAI into three classes (low, medium, and high), which were subsequently analyzed with linear regressions for micro-trend analysis.

ANOVA was performed to detect mean differences in DAI estimates within income groups: low (LIE), lower-middle (LMIE), upper-middle (UMIE), and high-income economies (HIE). For post-hoc analysis, we applied a Games-Howell test to adjust for variances and corrected the *p*-value via Holm adjustment (Supplementary Table 6, Supplementary Fig. 7). The goal was to examine the relationship between digital adoption and the suspected determinant of income level. We performed regression analyses and ANOVA using R software (version 4.0.3) ^68^.

### Explanation of findings

The review contextualized preceding findings with current literature and sought explanations for the relationship between DAI and COVID-19 values, guided by the PRISMA Extension for Scoping Reviews ^69^ (Supplementary Table 3). The search strategy combined alternative spellings of COVID-19 terms and pandemic-related keywords with “digit*” (Supplementary Note). This review covers English-language studies published in peer-reviewed scientific journals, including original research, review articles, and perspectives. Our search excluded news reports, comments, letters, policy briefs, and reports not fully accessible.

On May 3, 2021, we conducted a comprehensive literature search in MEDLINE/PubMed and Web of Science databases. We searched for publications published the day after the WHO China County Office was informed about case clusters in Wuhan, January 1, 2020 ^1^, to the day of screening. We first selected studies based on their titles and abstracts and then evaluated their full texts for inclusion. Duplicates were removed using EndNote.

Two authors (HH and LR) independently screened articles at both stages using Rayyan ^70^. Literature that did not meet the predefined criteria was removed. Disagreements between reviewers were resolved through discussion until inter-rater agreement was high (Cohen’s kappa coefficient >0.9). Our study team developed a screening manual with inclusion and exclusion criteria and a data extraction form (Supplementary Table 2 and Supplementary Table 4).

Eligible studies were thematically analyzed following Mays et al.’s narrative review approach ^71^. We identified and juxtaposed common keywords to capture knowledge from different sources. Recurrent keywords were identified by repeatedly examining the reviewed articles’ key pieces to ensure consistent interpretation and later merged them into themes. We mapped the keywords’ occurrences in a matrix, revealing considerable overlap between themes (Supplementary Table 5). The three themes are described as follows:

The first theme encompasses the importance of digital health tools integrated into the *health care system*. These include various electronic methods such as telemedicine to support clinical staff in managing patients or electronic medical records to facilitate the digital infrastructure of medical facilities and reduce massive pressures on hospitals. By strengthening health systems, hospital management, and clinical workflows, these technologies, in one way or another, may have a direct impact on decreasing morbidity and mortality.

The second theme refers to studies on *government responses* aiming at containing and mitigating the spread of COVID-19. For example, some studies focused on tracing and detecting suspected cases through comprehensive surveillance systems, digital contact tracing, and exposure notifications. Others addressed non-pharmaceutical interventions, including stay-at-home orders, restrictions of gatherings, and mask-wearing requirements. In addition, some studies have developed evidence-based guidance for policymakers and scientists in the form of epidemiologic models and empirical frameworks.

The third theme focused on literature involving papers that provide information on *determinants facilitating or hindering public health outcomes* beyond the themes mentioned above. Those papers revolve around risk communication and “infodemic” management on national, regional, and individual levels that are crucial in shaping individual health-seeking behavior. Next to communication, we found other factors that influenced pandemic management due to complex, interacting factors such as socioeconomic, organizational, political, and privacy concerns.

## Data Availability

All data produced in the present study are available upon reasonable request to the authors.

## Data Availability

The data that support the findings of this study are publicly available. We have compiled COVID-19 related data (i.e., number of cases and deaths and SI and demographic and epidemiological country-specific parameters) from *Our World in Data* at https://covid.ourworldindata.org/. The DAI data can be found at https://www.worldbank.org/en/publication/wdr2016/Digital-Adoption-Index and the World Bank data about countries’ income groups at https://datahelpdesk.worldbank.org/knowledgebase/articles/906519-world-bank-country-and-lending-groups. All reports selected for the scoping review and from which data were generated are included in the Supplementary Information.

## Acknowledgements

This work is funded by the German Federal Ministry of Education and Research (BMBF, grant no.: 01GP1910A).

## Author contributions

Conception and design: H.H. and F.K.; acquisition of data: H.H., F.M., and S.R.T.; analysis and interpretation of data: H.H., L.R., F.M., V.S., S.R.T., and F.K.; writing of the manuscript: H.H.; study supervision: F.K. All authors reviewed the manuscript and gave their final approval.

## Competing interests

All authors declare no competing interests.

## Additional information

Correspondence and requests for materials should be addressed to S.R.T. or F.K.

## Notes

### Competing Interest Statement

The authors have declared no competing interest.

### Funding Statement

This work has been funded by the German Federal Ministry of Education and Research (BMBF, grant no.: 01GP1910A).

### Summary of Updates

Figure 4 has been revised and the abstract and discussion updated accordingly.

