## Supplemental Information for "Digitalization impacts the COVID-19 pandemic and the stringency of government measures"

### Supplementary Methods

#### Country-specific examples for determining the decline in new deaths pm

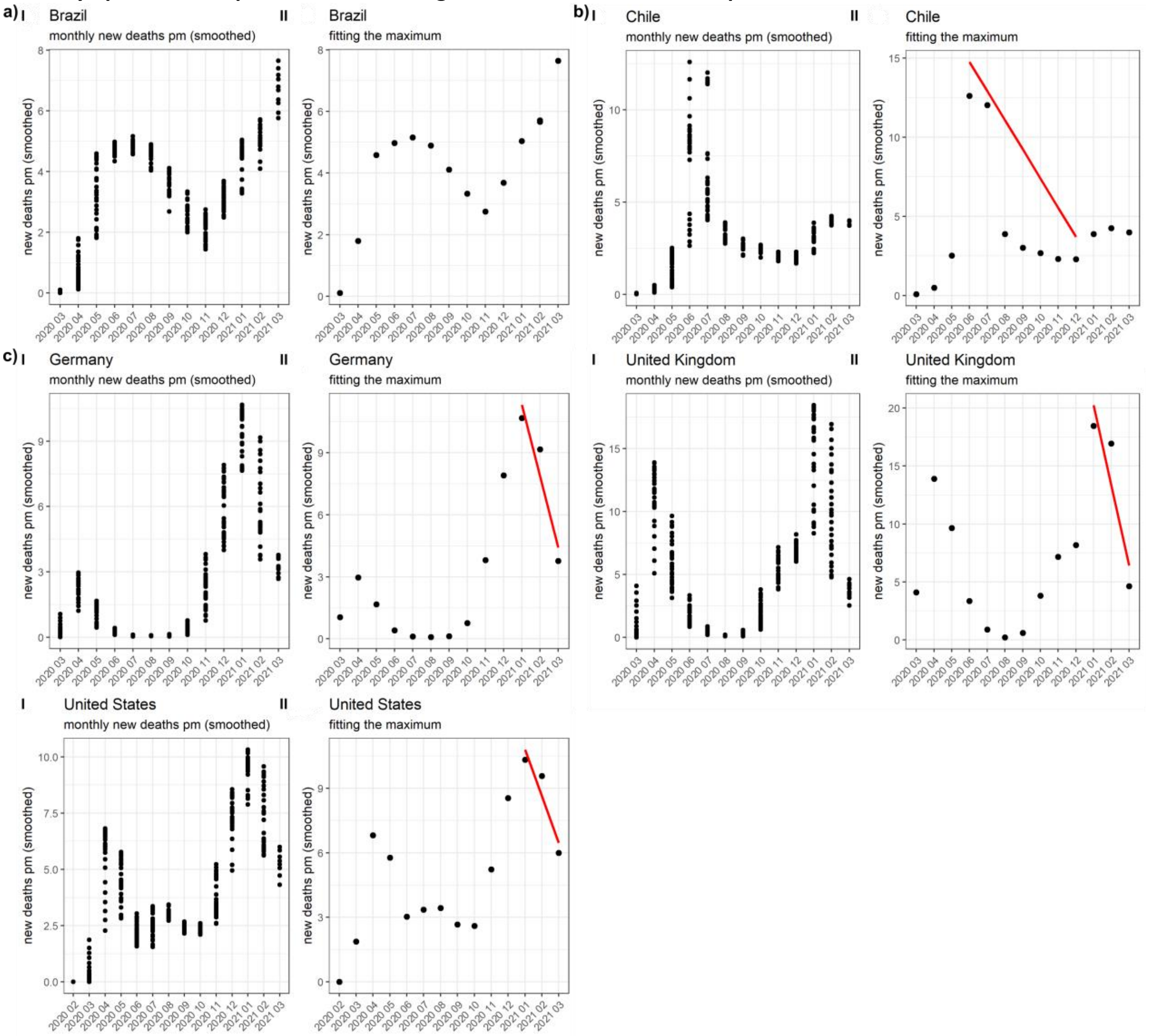

**Supplementary Figure 1.** Country-specific examples for determining the decline in new deaths pm in the post-peak period. The post-peak period describes the time and duration at which the spread of deaths has dropped below the peak. Linear regression was fit on the monthly data between the peak maximum and the maximum of the minimum that followed the peak. The period after the peak was determined separately for each country, and the slope values were finally included in the linear regression models. Countries were excluded if they a) were unable to reduce their death rates, b) experienced an increase in death rates that exceeded half of the peak height, or c) reached the peak at the end of the observation time (March 2021). Post-peak periods were analyzed for each country separately and, therefore, differed in length.

**Determining thresholds and subgroups in DAI**

Supplementary Table 1 presents the results of the treatment contrasts of the interaction of DAI and its three subgroups (low, medium, and high). This step is undertaken as an additional step after noticing three trends in data when plotting the $SI_{mean}$ against DAI. The DAI:subgroup interaction shows a significant effect (β=-109.44, $p<0.05$). Therefore, DAI slope is dependent on the subgroups. This also indicates that the effect of a country's digitalization depends on its relative position within the DAI scale.

**Supplementary Table 1.** Regression model of interaction effects of DAI and its subgroups.

| | β | Std. Error of β | t | *p-value* |
|---|---|---|---|---|
| ***Constant*** | 38.61 | 10.65 | 3.627 | 0.000** |
| **DAI** | 54.18 | 36.95 | 1.466 | 0.145 |
| **medium** | 32.56 | 14.28 | 2.280 | 0.024* |
| **high** | 63.74 | 27.23 | 2.341 | 0.021* |
| **DAI:medium** | -64.79 | 40.84 | -1.586 | 0.115 |
| **DAI:high** | -109.44 | 49.22 | -2.224 | 0.028* |

* $p \leq 0.05$; ** $p \leq 0.001$;

Notes: The regression coefficient (β) is the degree of change in the outcome variable for every one-unit change in the predictor variable. The t-statistic is the regression coefficient divided by its standard error. Digital adoption is expressed in arbitrary units of DAI. The *Constant* in this regression analysis represents the mean value of the low DAI subgroup (the intercept).

Supplementary Fig. 2 shows the subgroup thresholds [0.36; 0.68], obtained from the n=61 inferential interaction analyses with linear regression in the range DAI [0.2: 0.8]. The thresholds discriminate the changes in trend in the data and determine the three DAI subgroups (low, medium, and high). In the plot, each bubble represents the p-value from one of the interaction analyses. We considered the continuous flow of a $p \leq 0.05$ to code for the medium subgroup.

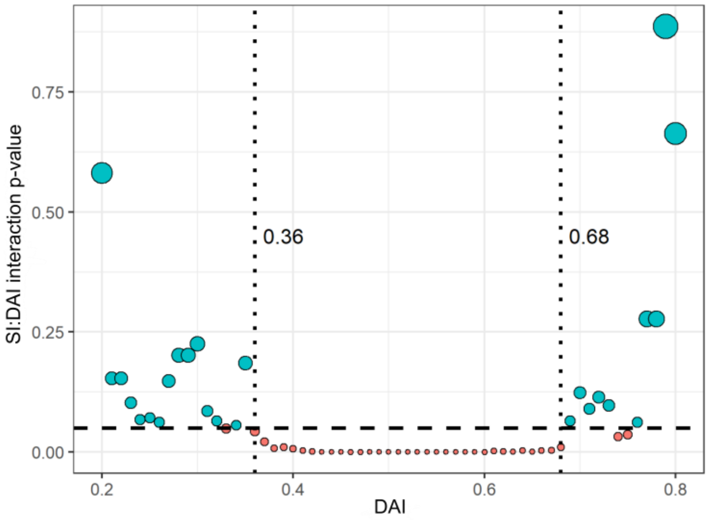

**Supplementary Figure 2.** Classification of thresholds in DAI. We observed three trends in DAI:SI interaction, where the p-value changed from $p \leq 0.05$ (red dots) to $p \geq 0.05$ (turquoise dots). The change in p-values determined the classification of the DAI into three categories (low, medium, and high). The dotted line shows the breaking points in data, respectively, thresholds of the three DAI categories [0.36; 0.68]. The different dot sizes represent the SI:DAI interaction p-values.

**Supplementary Note: Search strategy**

Search strategy used for PubMed (n=2,082).
("Coronavirus"[MeSH Terms] OR "Coronavirus"[Title/Abstract] OR "Coronaviruses"[Title/Abstract] OR "corona virus"[Title/Abstract] OR "sars-cov-2"[MeSH Terms] OR "sars-cov-2"[Title/Abstract] OR "Covid"[Title/Abstract] OR "Covid-19"[MeSH Terms] OR "Covid-19"[Title/Abstract] OR "Covid19"[Title/Abstract] OR "Coronaviridae"[MeSH Terms] OR "Coronaviridae"[Title/Abstract] OR "Coronavirinae"[Title/Abstract] OR "Betacoronavirus"[MeSH Terms] OR "Betacoronavirus"[Title/Abstract] OR "Betacoronaviruses"[Title/Abstract] OR "Ncov"[Title/Abstract] OR "pandemics"[MeSH Terms] OR "pandemic*"[Title/Abstract]) AND ("digit*"[Title/Abstract]) AND ("2020/01/01"[Date - Publication] : "3000"[Date - Publication])

Search strategy used for Web of Science (n=2,930).
((TS=(covid OR COVID-19 OR COVID19 OR coronavirus OR “corona virus” OR “coronavirus disease 2019” OR “SARS-CoV-2” OR nCoV OR pandemics OR coronaviridae OR coronavirinae OR betacoronavirus)) AND (TS=(digit*))) AND (DOP=(2020-01-01/2021-05-03))

### Eligibility criteria

**Supplementary Table 2.** Eligibility criteria. Inclusion and exclusion characteristics of the studies included in the scoping review.

| | |
|---|---|
| Type of publication and study design | - Studies published in a peer-reviewed scientific journal. We exclude articles that are in-press or pre-prints.<br>- Original research, reviews articles, and perspectives. We exclude news, rapid reports and communications, comments, conference abstracts, letters, policy briefs, and guidance, editorials, clinical studies, and articles where we failed to access full text. |
| Population | - All humans |
| Context | - The context of this paper will be global |
| Language restrictions | - All English-language papers |
| Publication date | - Articles published from 01/01/2020 to 03/05/2021 |
| Interventions, exposures | - All COVID-19 related (such as clinical features of COVID-19, investigations relating to COVID-19, outcomes relating to COVID-19, associations with COVID-19 incidence, mortality)<br>- Descriptions of and/or applications of digital health technology solutions in the context of COVID-19 OR in the context of population-level public health interventions (it may refer to interventions that were at stake before the pandemic started) |
| Outcomes | - Due to the broad nature of this scoping review, outcome measures analyzing any topic related to COVID-19 and its intersection with digitalization will be included.<br><br>This will include but is not limited to:<br>- COVID-19 outcomes, disease frequencies, and epidemiology (mortality, morbidity, incidence, prevalence, deaths, …)<br>- Digital technologies used in the COVID-19 pandemic<br>- Digital technologies used before the COVID-19 pandemic<br>- Administrative outcomes (government responses, infrastructure/organization of hospitals, utilization of digital technologies)<br>- Equipment/safety-related (intensive care units, access to hospitals and operating units)<br>- Countries' digital performance (access to digital technologies; adoption, preparedness, and readiness for digital development; digital infrastructure) |
| Sort and prioritize your exclusion criteria per selection phase (optional) | Selection phase: title/abstract screening<br>1. Not COVID-19 AND/OR digit* related<br>2. Not English<br>3. Wrong population<br>4. Wrong publication type/study design<br><br>Selection phase: full-text screening<br>1. No full text is accessible<br>2. Not (primarily) COVID-19 AND/OR digit* related<br>3. Off-topic: Articles that did primarily focus on digital tools for mitigating COVID-19<br>4. Wrong outcomes<br>5. Wrong publication type/study design |

**Preferred Reporting Items for Systematic Reviews and Meta-Analyses extension for Scoping Reviews**

**Supplementary Table 3.** Preferred Reporting Items for Systematic Reviews and Meta-Analyses extension for Scoping Reviews (PRISMA-ScR) Checklist.

| SECTION | ITEM | PRISMA-ScR CHECKLIST ITEM | REPORTED ON PAGE # |
|---|---|---|---|
| TITLE | | | |
| Title | 1 | Identify the report as a scoping review. | We did not include this information in the title, as the scoping review was one of three methodologies applied. |
| ABSTRACT | | | |
| Structured summary | 2 | Provide a structured summary that includes (as applicable): background, objectives, eligibility criteria, sources of evidence, charting methods, results, and conclusions that relate to the review questions and objectives. | 1 |
| INTRODUCTION | | | |
| Rationale | 3 | Describe the rationale for the review in the context of what is already known. Explain why the review questions/objectives lend themselves to a scoping review approach. | 2 |
| Objectives | 4 | Provide an explicit statement of the questions and objectives being addressed with reference to their key elements (e.g., population or participants, concepts, and context) or other relevant key elements used to conceptualize the review questions and/or objectives. | 12 |
| METHODS | | | |
| Protocol and registration | 5 | Indicate whether a review protocol exists; state if and where it can be accessed (e.g., a Web address); and if available, provide registration information, including the registration number. | n.a. |
| Eligibility criteria | 6 | Specify characteristics of the sources of evidence used as eligibility criteria (e.g., years considered, language, and publication status), and provide a rationale. | Supplementary Table 2 |
| Information sources* | 7 | Describe all information sources in the search (e.g., databases with dates of coverage and contact with authors to identify additional sources), as well as the date the most recent search was executed. | 12 |
| Search | 8 | Present the full electronic search strategy for at least 1 database, including any limits used, such that it could be repeated. | Supplementary Note |
| Selection of sources of evidence† | 9 | State the process for selecting sources of evidence (i.e., screening and eligibility) included in the scoping review. | Supplementary Table 2 |
| Data charting process‡ | 10 | Describe the methods of charting data from the included sources of evidence (e.g., calibrated forms or forms that have been tested by the team before their use, and whether data charting was done independently or in duplicate) and any processes for obtaining and confirming data from investigators. | 12-13 |
| Data items | 11 | List and define all variables for which data were sought and any assumptions and simplifications made. | Supplementary Table 4 |
| Critical appraisal of individual sources of evidence§ | 12 | If done, provide a rationale for conducting a critical appraisal of included sources of evidence; describe the methods used and how this information was used in any data synthesis (if appropriate). | n.a. |
| Synthesis of results | 13 | Describe the methods of handling and summarizing the data that were charted. | 12-13 |
| RESULTS | | | |
| Selection of sources of evidence | 14 | Give numbers of sources of evidence screened, assessed for eligibility, and included in the review, with reasons for exclusions at each stage, ideally using a flow diagram. | Supplementary Fig. 3 |

| SECTION | ITEM | PRISMA-ScR CHECKLIST ITEM | REPORTED ON PAGE # |
|---|---|---|---|
| Characteristics of sources of evidence | 15 | For each source of evidence, present characteristics for which data were charted and provide the citations. | Supplementary Table 3, Supplementary Fig. 4 & 5 |
| Critical appraisal within sources of evidence | 16 | If done, present data on critical appraisal of included sources of evidence (see item 12). | n.a. |
| Results of individual sources of evidence | 17 | For each included source of evidence, present the relevant data that were charted that relate to the review questions and objectives. | n.a. |
| Synthesis of results | 18 | Summarize and/or present the charting results related to the review questions and objectives. | Supplementary Table 3 |
| DISCUSSION | | | |
| Summary of evidence | 19 | Summarize the main results (including an overview of concepts, themes, and types of evidence available), link to the review questions and objectives, and consider the relevance to key groups. | 3-8 & Supplementary Results |
| Limitations | 20 | Discuss the limitations of the scoping review process. | 10 |
| Conclusions | 21 | Provide a general interpretation of the results with respect to the review questions and objectives, as well as potential implications and/or next steps. | 9-11 |
| FUNDING | | | |
| Funding | 22 | Describe sources of funding for the included sources of evidence, as well as sources of funding for the scoping review. Describe the role of the funders of the scoping review. | 19 |

JBI = Joanna Briggs Institute; PRISMA-ScR = Preferred Reporting Items for Systematic reviews and Meta-Analyses extension for Scoping Reviews.

* Where *sources of evidence* (see second footnote) are compiled from, such as bibliographic databases, social media platforms, and Web sites.

† A more inclusive/heterogeneous term used to account for the different types of evidence or data sources (e.g., quantitative and/or qualitative research, expert opinion, and policy documents) that may be eligible in a scoping review as opposed to only studies. This is not to be confused with *information sources* (see first footnote).

‡ The frameworks by Arksey and O'Malley (6) and Levac and colleagues (7) and the JBI guidance (4, 5) refer to the process of data extraction in a scoping review as data charting.

§ The process of systematically examining research evidence to assess its validity, results, and relevance before using it to inform a decision. This term is used for items 12 and 19 instead of "risk of bias" (which is more applicable to systematic reviews of interventions) to include and acknowledge the various sources of evidence that may be used in a scoping review (e.g., quantitative and/or qualitative research, expert opinion, and policy document).

*From:* Tricco AC, Lillie E, Zarin W, O'Brien KK, Colquhoun H, Levac D, et al. PRISMA Extension for Scoping Reviews (PRISMAScR): Checklist and Explanation. Ann Intern Med. 2018;169:467–473. doi: 10.7326/M18-0850.

### Supplementary Results

#### Flowchart of study selection process

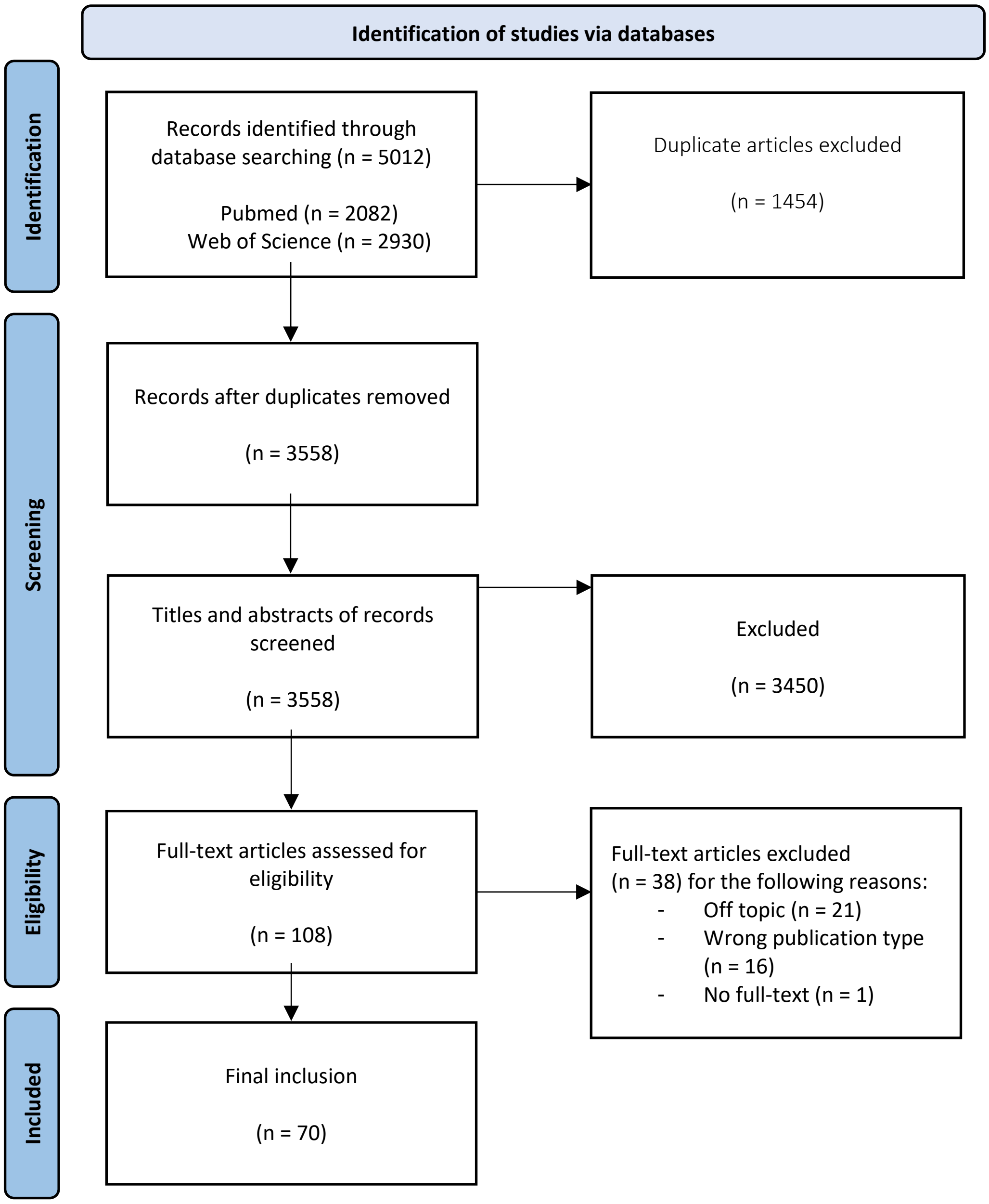

**Supplementary Figure 3.** Flowchart of screening and selection process.

### Data extraction table

**Supplementary Table 4.** List of included papers (n=70).

| First author | Publication Year | Title (type of article) | Description |
|---|---|---|---|
| Abdel-Basset, M. [1] | 2020 | An intelligent framework using disruptive technologies for COVID-19 analysis (original research) | The authors describe disruptive technologies and their benefits to combat COVID-19. |
| Abueg, M. & Hinch, R. [2] | 2021 | Modeling the effect of exposure notification and non-pharmaceutical interventions on COVID-19 transmission in Washington state (original research) | The influence of notifications combined with other non-pharmaceutical interventions to combat COVID-19 is analyzed. |
| Aggarwal, L. [3] | 2020 | Multi-criterion Intelligent Decision Support system for COVID-19 (original research) | Various technologies to combat COVID-19 in India are evaluated. |
| Alexiadis, A. [4] | 2020 | Simulation of pandemics in real cities: enhanced and accurate digital laboratories (original research) | The authors develop a framework to simulate the spread of infectious diseases in Birmingham and Bogotá. |
| Allam, M. & Cai, S. [5] | 2020 | COVID-19 Diagnostics, Tools, and Prevention (review article) | Recent trends in pandemic biology, diagnostics methods, prevention tools, and policies to combat COVID-19 are evaluated. |
| Almalki, A. [6] | 2021 | Health Apps for Combating COVID-19: Descriptive Review and Taxonomy (review article) | Mobile phone applications to combat COVID-19 are presented. |
| Anthony, B. [7] | 2021 | Integrating telemedicine to support digital health care for the management of COVID-19 pandemic (review article) | The authors discuss the benefits of telemedicine to combat COVID-19 worldwide. |
| Anthony, B. [8] | 2020 | Implications of telehealth and digital care solutions during COVID-19 pandemic: a qualitative literature review (review article) | The authors report on types of telehealth and digital care, policies, and the influence of telehealth and digital care solutions in countries worldwide. |
| Anttiroiki, A. V. [9] | 2021 | Successful Government Responses to the Pandemic: Contextualizing National and Urban Responses to the COVID-19 Outbreak in East and West (original research) | Various national strategies to combat COVID-19 are evaluated. |
| Asadzadeh, A. [10] | 2021 | A scope of mobile health solutions in COVID-19 pandemics (review article) | The authors analyze various mobile-health technologies to combat COVID-19. |
| Basellini, U. [11] | 2021 | Linking excess mortality to mobility data during the first wave of COVID-19 in England and Wales (original research) | The effects of mobility reductions in England and Wales between February and August 2020 are analyzed. |
| Behar, J. A. [12] | 2020 | Remote health diagnosis and monitoring in the time of COVID-19 (review article) | The authors evaluate remote health monitoring initiatives taken in 20 states. |
| Boeing, P. [13] | 2021 | Decoding China's COVID-19 'virus exceptionalism': Community-based digital contact tracing in Wuhan (original research) | The authors critically address China's exceptionalism in managing the COVID-19 pandemic with the example of the city Wuhan. |
| Budd, J. [14] | 2020 | Digital technologies in the public-health response to COVID-19 (review article) | Digital technologies to combat COVID-19 are analyzed in public health response to the pandemic. |
| Cencetti, G. & Santin, G. [15] | 2021 | Digital proximity tracing on empirical contact networks for pandemic control (original research) | The impact of digital contact tracing in combatting COVID-19 is evaluated using a framework based on contact data. |

| | | | |
|---|---|---|---|
| Chatterjee, P. [16] | 2020 | Internet of Things and Artificial Intelligence in Healthcare During COVID-19 Pandemic-A South American Perspective (review article) | The digital healthcare in South America during the COVID-19 pandemic is discussed. |
| Chaudhary, Y. [17] | 2021 | Digital Warfare Against COVID-19: Global Use of Contact-Tracing Apps (review article) | Contact tracing apps worldwide are evaluated regarding their safety, privacy, and efficacy. |
| Elavarasan, R. M. [18] | 2021 | A hover view over effectual approaches on pandemic management for sustainable cities - The endowment of prospective technologies with revitalization strategies (original research) | The authors analyze the characteristics of the pandemic as well as the role of technologies to manage it and lessons learnt. |
| Faraj, S. [19] | 2021 | Unto the breach: What the COVID-19 pandemic exposes about digitalization (original research) | Data are presented on how the COVID-19 pandemic challenges digitalization issues. |
| Fariniuk, T. M. D. [20] | 2020 | Smart cities and the pandemic: digital technologies on the urban management of Brazilian cities (original research) | The use of digital tools in Brazilian cities to combat COVID-19 is explored. |
| Ferretti, L. & Wymant, C. [21] | 2020 | Quantifying SARS-CoV-2 transmission suggests epidemic control with digital contact tracing (original research) | Various transmission routes of COVID-19 were investigated. |
| Golinelli, D. & Boetto, E. [22] | 2020 | Adoption of Digital Technologies in Health Care During the COVID-19 Pandemic: Systematic Review of Early Scientific Literature (review article) | Digital solutions to mitigate the impact of COVID-19 on individuals and the health systems are evaluated. |
| Gunasekeran, D. V. & Tseng, R. M. W. W. [23] | 2021 | Applications of digital health for public health responses to COVID-19: a systematic scoping review of artificial intelligence, telehealth and related technologies (review article) | Digital health solutions for public health responses are discussed. |
| He, Z., Zhang, C. J. P. & Huang, J. [24] | 2020 | A New Era of Epidemiology: Digital Epidemiology for Investigating the COVID-19 Outbreak in China (perspective article) | In this viewpoint, the authors estimate the disease spread patterns related to human activities. |
| Heo, K. [25] | 2020 | Searching for Digital Technologies in Containment and Mitigation Strategies: Experience from South Korea COVID-19 (review article) | Field applications of governmental interventions to combat COVID-19 are investigated. |
| Hernández-Orallo, E. [26] | 2020 | Evaluating the Effectiveness of COVID-19 Bluetooth-Based Smartphone Contact Tracing Applications (original research) | Newly developed contact tracing mobile applications to combat COVID-19 are evaluated. |
| Hernández-Orallo, E. [27] | 2020 | Evaluating How Smartphone Contact Tracing Technology Can Reduce the Spread of Infectious Diseases: The Case of COVID-19 (original research) | Contact tracing technologies and their impact on controlling infectious diseases are studied. |
| Huang, Z. [28] | 2020 | Performance of Digital Contact Tracing Tools for COVID-19 Response in Singapore: Cross-Sectional Study (original research) | The authors analyze the performance of the contact tracing app "Trace Together" and compares it to a wearable tag-based real-time locating system. |
| Islam, M. N. [29] | 2020 | A Systematic Review of the Digital Interventions for Fighting COVID-19: The Bangladesh Perspective (review article) | The authors analyze digital interventions to combat COVID-19 in Bangladesh and compare them to other countries. |
| Javaid, M. [30] | 2020 | Industry 4.0 technologies and their applications in fighting COVID-19 pandemic (review article) | Technologies of Industry 4.0 and their applications to combat COVID-19 are evaluated. |

| | | | |
|---|---|---|---|
| Javaid, M. [31] | 2020 | Industry 5.0: Potential Applications in COVID-19 (review article) | Industry 5.0 technologies that can be used to combat COVID-19 are explored. |
| Jian, S. W. [32] | 2020 | Contact tracing with digital assistance in Taiwan's COVID-19 outbreak response (perspective article) | The authors developed a centralized contact tracing system to combat COVID-19. |
| Jung, S. Y. [33] | 2020 | How IT preparedness helped to create a digital field hospital to care for COVID-19 patients in S. Korea (perspective article) | The impact of IT preparedness on creating a digital field hospital in South Korea is analyzed. |
| Kalhori, S. R. N. [34] | 2021 | Digital Health Solutions to Control the COVID-19 Pandemic in Countries With High Disease Prevalence: Literature Review (review article) | The application of digital technologies to combat COVID-19 in the ten countries with the highest prevalence of the disease is investigated. |
| Khan, Z. H. [35] | 2020 | Robotics Utilization for Healthcare Digitization in Global COVID-19 Management (original research) | The role of robotics in healthcare regarding the mastery of COVID-19 is studied. |
| Kogan, N. E., Clemente, L. & Liautaud, P. [36] | 2021 | An early warning approach to monitor COVID-19 activity with multiple digital traces in near real time (original research) | Digital data streams are analyzed regarding their role as indicators of COVID-19 activity. |
| Król, K. [37] | 2021 | The Quality of Infectious Disease Hospital Websites in Poland in Light of the COVID-19 Pandemic (original research) | The authors evaluate hospital websites in Poland in matters regarding COVID-19. |
| Kucharski, A. J. [38] | 2020 | Effectiveness of isolation, testing, contact tracing, and physical distancing on reducing transmission of SARS-CoV-2 in different settings: a mathematical modelling study | The effectiveness of COVID-19 transmission reduction strategies is estimated. |
| Lai, S. H. S. [39] | 2020 | The experience of contact tracing in Singapore in the control of COVID-19: highlighting the use of digital technology (review article) | Forms of contact tracing, especially new (digital) technology innovations used in Singapore to combat COVID-19, are evaluated. |
| Leung, K. & Wu, J. T. [40] | 2021 | Real-time tracking and prediction of COVID-19 infection using digital proxies of population mobility and mixing (original research) | The authors develop a framework that quantifies disease transmission models with age-specific digital mobility data. |
| Liaw, S. T. [41] | 2021 | Primary Care Informatics Response to Covid-19 Pandemic: Adaptation, Progress, and Lessons from Four Countries with High ICT Development (review article) | The authors evaluate the response to COVID-19 in Australia, Canada, the United Kingdom and the United States regarding primary care informatics. |
| Lin, L. [42] | 2020 | Combat COVID-19 with artificial intelligence and big data (perspective article) | The use of artificial intelligence and big data to combat COVID-19 in China (Mainland and Hongkong), Taiwan, and South Korea are evaluated. |
| Mahdi, A. [43] | 2021 | OxCOVID19 Database, a multimodal data repository for better understanding the global impact of COVID-19 (original research) | The Oxford COVID-19 Database is described. |
| Martin, T. [44] | 2020 | Demystifying COVID-19 Digital Contact Tracing: A Survey on Frameworks and Mobile Apps (review article) | The authors analyze the most concrete contact tracing architectures worldwide. |
| Meijer, A. [45] | 2020 | The COVID-19-crisis and the information polity: An overview of responses and discussions in twenty-one countries from six continents (review article) | The governmental responses to COVID-19 in 21 countries are compared, especially regarding their information policy. |

| | | | |
|---|---|---|---|
| Nachega, J. B. [46] | 2021 | Contact Tracing and the COVID-19 Response in Africa: Best Practices, Key Challenges, and Lessons Learned from Nigeria, Rwanda, South Africa, and Uganda (perspective article) | The COVID-19 contact tracing in Nigeria, Rwanda, South Africa and Uganda is evaluated. |
| Nageshwaran, G. [47] | 2021 | Review of the role of big data and digital technologies in controlling COVID-19 in Asia: Public health interest vs. privacy (review article) | The authors evaluate the digital tools used to combat COVID-19 in Taiwan, South Korea, China and Singapore. |
| Nuzzo, A. [48] | 2020 | Universal Shelter-in-Place Versus Advanced Automated Contact Tracing and Targeted Isolation: A Case for 21st-Century Technologies for SARS-CoV-2 and Future Pandemics (original research) | Digital contact tracing and shelter-in-place actions regarding their effect on the spread of COVID-19 are discussed. |
| O'Connor, H. [49] | 2021 | For the greater good? Data and disasters in a post-COVID world (research article) | The difficulties of data usage during the COVID-19 pandemic in Taiwan and New Zealand are addressed. |
| Reddick, C. G. [50] | 2020 | Determinants of broadband access and affordability: An analysis of a community survey on the digital divide (original research) | The digital divide is analyzed in a case study in San Antonio. |
| Rodríguez, P. [51] | 2021 | A population-based controlled experiment assessing the epidemiological impact of digital contact tracing (original research) | The authors conducted a population-based controlled experiment in La Gomera (Canary Islands) to analyze the impact of the Radar COVID mobile application. |
| Salathé, M. [52] | 2020 | Early evidence of effectiveness of digital contact tracing for SARS-CoV-2 in Switzerland (original research) | Digital contact tracing applications available in Switzerland are reported. |
| Schlosser, F. [53] | 2020 | COVID-19 lockdown induces disease-mitigating structural changes in mobility networks (original research) | The structural mobility changes during the COVID-19 pandemic in Germany are analyzed. |
| Scott, B. K. [54] | 2020 | Advanced Digital Health Technologies for COVID-19 and Future Emergencies (review article) | The authors evaluate digital health technologies useful to combat COVID-19. |
| Shamil, M. S. [55] | 2020 | An Agent-Based Modeling of COVID-19: Validation, Analysis, and Recommendations (research article) | The authors use an agent-based model to simulate the spread of COVID-19. |
| Skoll, D. [56] | 2020 | COVID-19 testing and infection surveillance: Is a combined digital contact-tracing and mass-testing solution feasible in the United States? (review article) | Technology-augmented contact-based surveillance is analyzed regarding their role in monitoring the COVID-19 outbreak. |
| Storeng, K. T. [57] | 2021 | The Smartphone Pandemic: How Big Tech and public health authorities' partner in the digital response to Covid-19 (review article) | The authors critically discuss new forms of public-private cooperation caused by the COVID-19 pandemic. |
| Sullivan, C. [58] | 2021 | Moving Faster than the COVID-19 Pandemic: The Rapid, Digital Transformation of a Public Health System (original research) | In this case study, the digitization of the public health system in Queensland, Australia, is analyzed regarding its benefits and lessons learnt during the COVID-19 pandemic. |
| Tejedor, S. [59] | 2020 | Tracking Websites' Digital Communication Strategies in Latin American Hospitals During the COVID-19 Pandemic (original research) | The highest-ranked hospitals in Latin America are analyzed regarding their websites during the COVID-19 pandemic. |
| Thomas, M. J. [60] | 2021 | Can technological advancements help to alleviate COVID-19 pandemic? a review (review article) | This review provides technological solutions which can be useful in combatting COVID-19. |

| | | | |
|---|---|---|---|
| Tiirinki, H [61] | 2020 | COVID-19 pandemic in Finland - Preliminary analysis on health system response and economic consequences (research article) | The authors analyze the effects of COVID-19 in Finland regarding health policy, the social- and health system and the economic system. |
| Whitelaw, S. [62] | 2020 | Applications of digital technology in COVID-19 pandemic planning and response (perspective article) | In this viewpoint, the authors describe a framework for applying digital technologies to combat COVID-19 and analyze how various countries have adopted these technologies. |
| Wood, A. [63] | 2021 | Linked electronic health records for research on a nationwide cohort of more than 54 million people in England: data resource (original research) | An electronic health record resource and its usefulness in fighting the COVID-19 pandemic is described. |
| Xu, X., Cai, Y. & Wu, S. [64] | 2021 | Assessment of Internet Hospitals in China During the COVID-19 Pandemic: National Cross-Sectional Data Analysis Study (original research) | The authors analyze China's internet hospitals and their health service capacity. |
| Yan, A. [65] | 2020 | How hospitals in mainland China responded to the outbreak of COVID-19 using information technology-enabled services: An analysis of hospital news webpages (original research) | The application and impact of information technology on Chinese hospitals are investigated. |
| Ye, Q. & Zhou, J. [66] | 2020 | Using Information Technology to Manage the COVID-19 Pandemic: Development of a Technical Framework Based on Practical Experience in China (original research) | The authors develop a framework to respond to COVID-19 from a health informatics perspective. |
| Yen, W. T. [67] | 2020 | Taiwan's COVID-19 Management: Developmental State, Digital Governance, and State-Society Synergy (review article) | In this review of policy practice, the authors analyze Taiwan's response strategy to COVID-19. |
| Zanin, M. [68] | 2020 | The public health response to the COVID-19 outbreak in mainland China: a narrative review (review article) | The authors analyze China's response strategy to COVID-19. |
| Zeng, K. & Bernardo, S. N. [69] | 2020 | The Use of Digital Tools to Mitigate the COVID-19 Pandemic: Comparative Retrospective Study of Six Countries (original research) | Precautions to combat COVID-19 in various countries are compared. |
| Zhao, Z. [70] | 2021 | Applications of Robotics, Artificial Intelligence, and Digital Technologies During COVID-19: A Review (review article) | News articles and scientific reports on technology adoption in combatting COVID-19 are analyzed. |

### Distribution of authors' residence

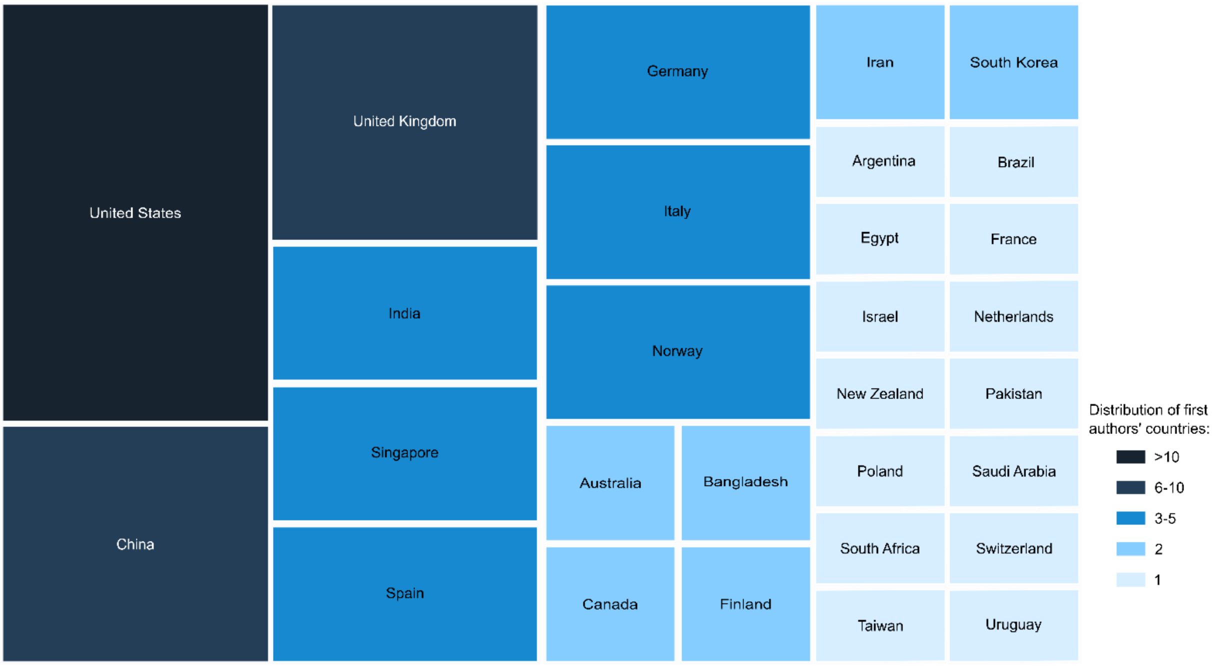

**Supplementary Figure 4.** The distribution of first authors' residence. This also includes multiple countries in the case of co-first authorship.

### Distribution of countries evaluated in the included papers

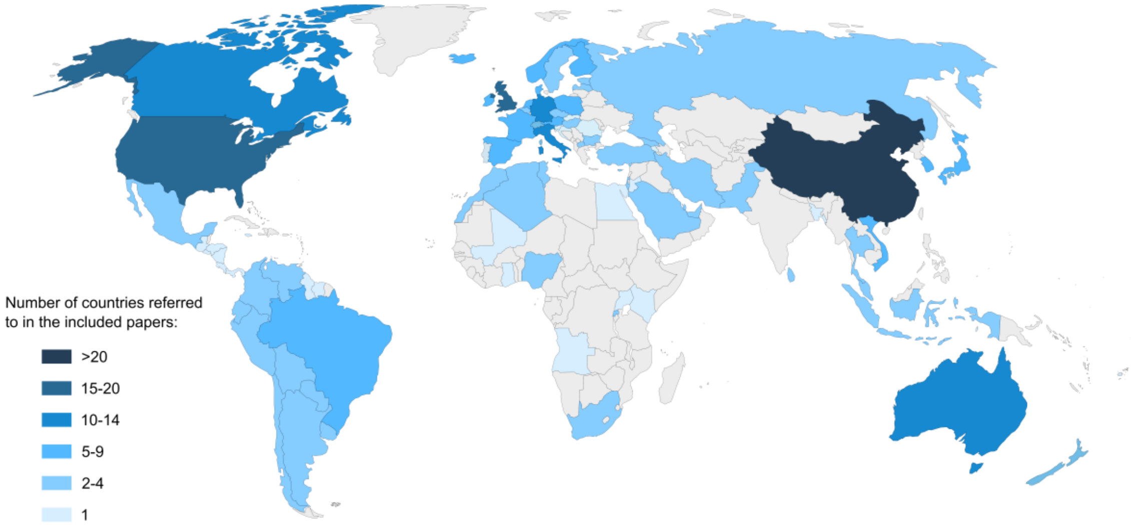

**Supplementary Figure 5.** Geographical map of screened reports. The distribution of countries evaluated in articles included in the scoping review.

### Data synthesis

**Supplementary Table 5.** Data synthesis. This matrix illustrates the occurrence of keywords and themes for each reference included in this scoping review.

| | | Healthcare system | | | Government response | | | Determinants of public health outcomes | | |
|---|---|---|---|---|---|---|---|---|---|---|
| **No.** | **First author** | **digital infrastructure** | **clinical managment** | **telehealth** | **contact tracing** | **non-pharmaceutical interventionss** | **evidence-based guidance** | **risk communication** | **sociocultural & political context** | **public attitudes** |
| 1 | Abdel-Basset, M. | 1 | | | | | 1 | | | |
| 2 | Abueg, M. & Hinch, R. | | | | 1 | 1 | | | | |
| 3 | Aggarwal, L. | | | | | | 1 | | | |
| 4 | Alexiadis, A. | | | | | | 1 | | | |
| 5 | Allam, M. | | | | 1 | | 1 | | | |
| 6 | Almalki, A. | | | | 1 | | | | | |
| 7 | Anthony, B. | | 1 | 1 | | | | | | |
| 8 | Anthony, B. | 1 | 1 | 1 | | | | | 1 | |
| 9 | Anttiroiki, A. V. | | | | | 1 | | | 1 | 1 |
| 10 | Asadzadeh, A. | | | 1 | 1 | | | | | |
| 11 | Basellini, U. | | | | 1 | 1 | | | | |
| 12 | Behar, J. A. | 1 | | 1 | | | | | | 1 |
| 13 | Boeing, P. | | | | 1 | | | | | |
| 14 | Budd, J. | | | | 1 | | | 1 | 1 | 1 |
| 15 | Cencetti, G. & Santin, G. | | | | 1 | 1 | 1 | | | |
| 16 | Chatterjee, P. | 1 | 1 | 1 | | | | | | |
| 17 | Chaudhary, Y. | | | | 1 | | | | | |
| 18 | Elavarasan, R. M. | | | | | | 1 | | | |
| 19 | Faraj, S. | | | | | | | | 1 | 1 |
| 20 | Fariniuk, T. M. D. | | | | | | | | 1 | |
| 21 | Ferretti, L. & Wymant, C. | | | | 1 | 1 | | | | |
| 22 | Golinelli, D. | 1 | 1 | 1 | | | | | | |
| 23 | Gunasekeran, D. V. | 1 | | 1 | | | | | | |
| 24 | He, Z., Zhang, C. J. P., and Huang, J. | | | | 1 | 1 | 1 | | | |
| 25 | Heo, K. | | | | | 1 | | | | |
| 26 | Hernández-Orallo, E. | | | | 1 | | 1 | | | |
| 27 | Hernández-Orallo, E. | | | | 1 | | 1 | | | |
| 28 | Huang, Z. | 1 | | | 1 | | | | | |
| 29 | Islam, M. N., and Islam, A. K. M. N. | | | | | 1 | | | | |
| 30 | Javaid, M. | 1 | | 1 | | | | | | |

| | | | | | | | | | | |
|---|---|---|---|---|---|---|---|---|---|---|
| 31 | Javaid, M. | 1 | 1 | 1 | | | | | | |
| 32 | Jian, S. W. | | | | 1 | | | | | |
| 33 | Jung, S. Y. | 1 | 1 | 1 | | | | | | |
| 34 | Kalhori, S. R. N. | 1 | | 1 | | | | | | |
| 35 | Khan, Z. H. | 1 | 1 | | | | | | | |
| 36 | Kogan, N. | | | | | 1 | | | | |
| 37 | Król, K. | 1 | | | | | | 1 | | |
| 38 | Kucharski, A. J. | | | | 1 | 1 | 1 | | | |
| 39 | Lai, S. H. S. | | | | 1 | | | | | |
| 40 | Leung, K. | | | | | | 1 | | | |
| 41 | Liaw, S. T. | 1 | 1 | 1 | | | | | 1 | |
| 42 | Lin, L. | | | | 1 | 1 | | | | |
| 43 | Mahdi, A. | | | | | | 1 | | | |
| 44 | Martin, T. | | | | 1 | | 1 | | | |
| 45 | Meijer, A. | | 1 | | 1 | 1 | | 1 | 1 | |
| 46 | Nachega, J. B. | | | | 1 | | | | | |
| 47 | Nageshwaran, G. | | | | 1 | 1 | | | | |
| 48 | Nuzzo, A. | | | | 1 | 1 | | | | |
| 49 | O'Connor, H. | | | | 1 | 1 | | | | 1 |
| 50 | Reddick, C. G. | | | | | | | | 1 | |
| 51 | Rodríguez, P. | | | | 1 | | | | | |
| 52 | Salathé, M. | | | | 1 | | | | | |
| 53 | Schlosser, F. | | | | | 1 | | | | |
| 54 | Scott, B. K. | 1 | 1 | 1 | | | | | | |
| 55 | Shamil, M. S. | | | | 1 | 1 | 1 | | | |
| 56 | Skoll, D. | | | | 1 | 1 | | | | 1 |
| 57 | Storeng, K. T. | | | | 1 | | | 1 | | 1 |
| 58 | Sullivan, C. | 1 | | | | | | | | |
| 59 | Tejedor, S. | 1 | | | | | | 1 | | |
| 60 | Thomas, M. J. | | | | | | | 1 | | |
| 61 | Tiirinki, H. | | | | | 1 | | | 1 | |
| 62 | Whitelaw, S. | 1 | 1 | 1 | 1 | 1 | | | 1 | |
| 63 | Wood, A. | 1 | | | | | | | | |
| 64 | Xu, X. | 1 | | 1 | | | | | | |
| 65 | Yan, A. | 1 | 1 | 1 | | | | 1 | | |
| 66 | Ye, Q. | 1 | | | | | 1 | | | |
| 67 | Yen, W. T. | | | | | 1 | | 1 | 1 | 1 |
| 68 | Zanin, M. | | | | 1 | 1 | | | | |
| 69 | Zeng, K. | | | | 1 | 1 | | | | |
| 70 | Zhao, Z. | 1 | 1 | 1 | | | | | | |
| | **Totals** | 23 | 13 | 17 | 32 | 23 | 15 | 8 | 11 | 8 |

Note: Totals reported in the results section of the manuscript correspond to the number of papers to which at least one of the three keywords could be assigned per topic: i) healthcare system (n=26), ii) government responses (n=46), and iii) determinants facilitating or hindering public health outcomes (n=19).

**Literature findings of the scoping review: Determinants facilitating or hindering public health outcomes**

For the final theme of our scoping review, we combined the literature discussing influential factors that hindered or facilitated the early response to the pandemic and that do not directly relate to digital technologies used in clinical settings or by governments. Three keywords emerged under this theme: socioeconomic and political contexts, risk communication, and public attitudes. Supplementary Fig. 3 shows the occurrence and overlap of all nine keywords included in the scoping review. Large disparities in affordability and access to mobile communications widen the digital divide [14]. Low-income settings, ethnic minorities, less educated groups, and the digitally illiterate are disproportionally affected by digital inequalities [50]. Reddick et al. illustrate this with a case study of broadband access in San Antonio, USA, where more than half the population is Hispanic. Residents of poorer neighborhoods had significantly lower rates of broadband use, defined here as an essential factor in a person's quality of life. This study highlights the significant digital divide within geographic structures [50]. Anthony et al. identified barriers to adopting digital health solutions at the infrastructural and institutional level, similar to the DAI pillars. They ultimately created determinants meaningful in adopting telehealth [8]. In addition, great exposure to information, inappropriate risk communication, or even misinformation have confused and influenced individual health behaviors. Misinformation and changing guidance have led people to reject public health measures such as wearing masks and physical distancing [14,60]. Finally, digital technologies raise legal, ethical, and privacy concerns [8,57].

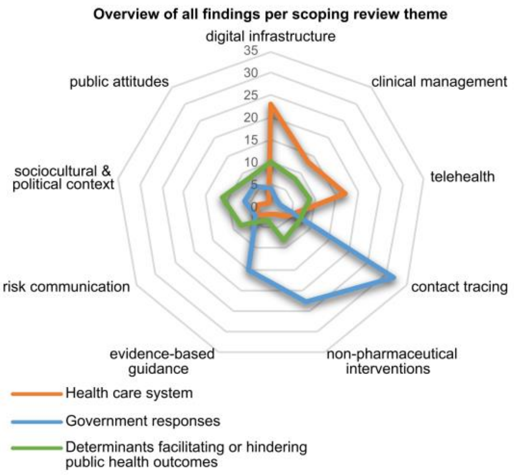

**Supplementary Figure 6.** Occurrence of keywords and themes within the scoping review. The radar chart illustrates the frequency and overlap of studies assigned to the following three themes of the scoping review: i) health care system, ii) government responses, and iii) determinants facilitating or hindering public health outcomes. The results of the first two themes are extensively described in the main manuscript. The third theme (green) includes keywords from all three themes. The axis of the radar chart represents the total number of included keywords within the themes (see Supplementary Table 5). The scale for the radial aces of this chart is the total number of reports per keyword.

### Global level of digital adoption by income

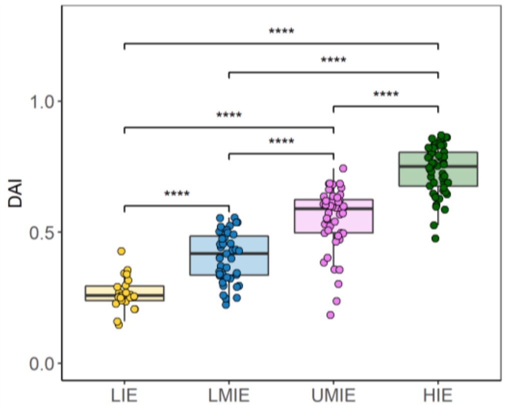

**Supplementary Figure 7.** Distribution of digital adoption by income group. Box and Whisker plot of the digital adoption index (DAI) by income level: low, lower-middle, upper-middle, and high income. One-way ANOVA of income groups followed by a Games-Howell post-hoc test for pairwise comparisons. DAI showed statistically significant differences between all groups ($p<0{\cdot}001$). (Note: LIE = low-income economy; LMIE = lower-middle-income economy; UMIE = upper-middle-income economy; HIE = high-income economy).

**Supplementary Table 6.** Post-hoc comparison of means between income groups.

| group 1 | group 2 | t | p-value (adj.) |
|---|---|---|---|
| HIE | LIE | -36.532 | 0.0001* |
| HIE | LMIE | -24.341 | 0.0001* |
| HIE | UMIE | -11.965 | 0.0001* |
| LIE | LMIE | 10.246 | 0.0001* |
| LIE | UMIE | 18.187 | 0.0001* |
| LMIE | UMIE | 8.889 | 0.0001* |

* $p<0.0001$
